# A non-muscle myosin heavy chain 9 genetic variant is associated with graft failure following kidney transplantation

**DOI:** 10.1101/2022.03.29.22272996

**Authors:** Felix Poppelaars, Siawosh K. Eskandari, Jeffrey Damman, Marc A. Seelen, Bernardo Faria, Mariana Gaya da Costa

## Abstract

**Background:** Despite current matching efforts to identify optimal donor-recipient pairs in kidney transplantation, alloimmunity remains a major proponent of late transplant failure. While kidney allocation based on human leukocyte antigen (HLA) matching has markedly prolonged short-term graft survival, new data suggests that additional genetic parameters in donor-recipient matching could help improve the long-term outcomes. Here, we studied the impact of a recently discovered non-muscle myosin heavy chain 9 gene (*MYH9*) polymorphism on kidney allograft failure.

**Methods:** We conducted a prospective observational cohort study, analyzing the DNA of 1,271 kidney donor-recipient transplant pairs from a single academic hospital for the *MYH9* rs11089788 C>A polymorphism. The association of the *MYH9* genotype with the risk of graft failure (primary outcome), biopsy-proven acute rejection (BPAR), and delayed graft function (DGF) (secondary outcomes) were determined.

**Results:** The *MYH9* polymorphism in the donor was not associated with 15-year death-censored kidney graft survival, whereas a trend was seen for the association between the *MYH9* polymorphism in the recipient and graft failure (recessive model, *P*=0.056). Having the AA-genotype of the *MYH9* polymorphism in recipients was associated with a higher risk of DGF (*P*=0.031) and BPAR (*P*=0.021), although the significance was lost after adjustment for potential confounders (*P*=0.15 and *P*=0.10, respectively). The combined presence of the *MYH9* polymorphism in donor-recipient pairs was significantly associated with long-term kidney allograft survival (*P*=0.036), in which recipients with an AA-genotype receiving a graft with an AA-genotype had the worst outcome. After adjustment for covariates, this combined genotype remained significantly associated with 15-year death-censored kidney graft survival (HR 1.68, 95%-CI: 1.05 – 2.70, *P*=0.031).

**Conclusions:** Our results reveal that recipients with an AA-genotype *MYH9* polymorphism receiving a donor kidney with an AA-genotype, have a significantly elevated risk of graft failure after kidney transplantation.

**Key points:**

- In recipients, the *MYH9* SNP was associated with delayed graft function and biopsy-proven acute rejection after kidney transplantation, although the significance was lost in multivariable analysis.
- Presence of the *MYH9* variant in both the donor and recipient significantly associated with long-term kidney allograft survival in multivariable analysis.
- Our present findings suggests that matching donor-recipient transplant pairs based on the *MYH9* polymorphism may attenuate the risk of graft loss.

## Introduction

Despite the excellent short-term outcomes following solid organ transplantation, the long-term survival of kidney transplants has only negligibly improved in recent years.^1^ As a consequence, one out of five patients on the waitlist for kidney transplantation are candidates whose previous grafts failed.^2^ Maximizing the long-term outcomes of transplantation and preventing re-transplantation is, thus, paramount—not only for improving transplant recipients’ outcomes, but also for reducing waitlist pressures. Among the many drivers of late graft loss, alloimmunity, otherwise known as host anti-donor immune responses, remains to be the preeminent cause, notably, despite efforts to optimally match donor-recipient pairs.^3,4^ Recently, there are signs of a paradigm shift in the transplant field, with suggestions that allograft matching efforts should be updated to include novel genetic markers that better ensure long-term graft survival after kidney transplantation.^5,6^

In this regard, non-muscle myosin heavy chain II-A (MHCII-A), encoded by the myosin heavy chain 9 gene (*MYH9*), is a target of particular interest (Fig. 1A). Non-muscle MHCII-A is a ubiquitously expressed contractile protein involved in a myriad of processes ranging from cell division and adhesion to providing cytoskeletal support.^7^ Mutations in the *MYH9* cause a complex set of disorders, known as *MYH9*-related diseases, that can affect every system in the body but are characterized by congenital thrombocytopenia, giant platelets and leucocyte inclusions.^7^ Although non-muscle MHCII-A is expressed by a variety of cell types, the podocyte lineage in particular, expressed high levels of this protein.^7^ Unsurprisingly, patients with *MYH9*-related disorders can clinically present with persistent proteinuria and a progressive decline in kidney function leading to end-stage kidney disease (ESKD).^7,8^ Subsequent studies linked common *MYH9* polymorphisms to an increased risk of developing focal segmental glomerulosclerosis and non-diabetic ESKD.^9,10^ However, it is worthy to note that these associations were later shown to be dependent on a strong linkage disequilibrium of these *MYH9* polymorphisms with variants in the apolipoprotein L1 gene (*APOL1*).^7,11^ Still, there are studies that show an association between *MYH9* polymorphisms and chronic kidney disease (CKD) independently of linkage with *APOL1*, suggesting a potential role for *MYH9* polymorphisms in the pathogenesis of ESKD.^12,13^

**Figure 1.**
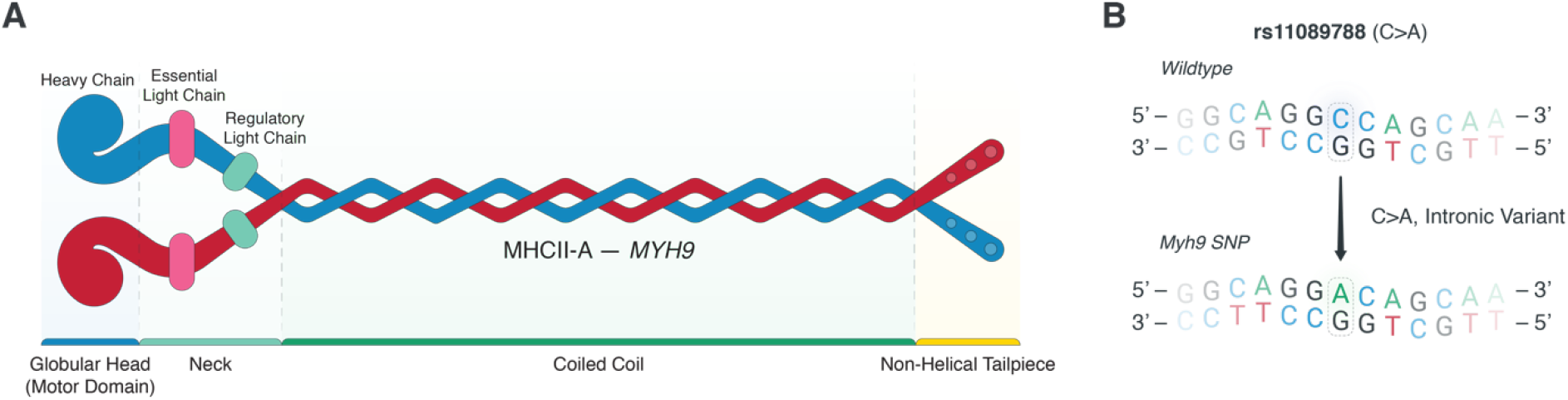
Illustration of the non-muscle myosin heavy chain 9 and the examined *MYH9* polymorphisms. (**A**) Non-muscle myosin heavy chain II-A (MHCII-A) is a contractile protein comprised of several domains: A globular motor head portion (heavy chain), a neck domain (essential light chain and regulatory light chain), coiled coil tail segment (MHCII-A), and non-helical tailpiece that can be phosphorylated. The coiled coil segment is notably encoded by the myosin heavy chain 9 (*MYH9*) gene. (**B**) In this study, we assessed the association of rs11089788 (C>A) *MYH9* single-nucleotide polymorphism (SNPs) in kidney allograft donors and recipients with long-term graft survival outcomes. MHCII-A, non-muscle heavy chain II-A; *MYH9*, myosin heavy chain 9 gene; SNP, single nucleotide polymorphism.

In a recent genome-wide linkage analysis, a significant association between the *MYH9* rs11089788 polymorphism and kidney function was identified in a meta-analysis of three European populations.^14^ This *MYH9* polymorphism was additionally found to be significantly associated with progressive loss of kidney function in other cohorts.^13,15^ Importantly, the associations between *MYH9* rs11089788 and kidney function could not be explained by linkage disequilibrium with *APOL1*.^15^

Here, we investigated the impact of the recently discovered rs11089788 *MYH9* polymorphism on long-term graft survival in the context of kidney transplantation (Fig. 1B). As a secondary outcome, we also assessed the association of this polymorphism with biopsy-proven acute rejection (BPAR) and delayed graft function (DGF).

## Materials and Methods

### Patient selection and study end-point

Patients receiving a single kidney transplantation at the University Medical Center Groningen (UMCG) in the Netherlands were recruited between March 1993 and February 2008. 1,271 of the 1,430 screened donor-recipient kidney transplant pairs were included in the current study as previously reported.^16–21^ Ethical approval for this study and the study protocol was given by the Institutional Review Board of the University Medical Center Groningen in Groningen, The Netherlands (Medical Ethical Committee 2014/077). The study protocol adhered to the Declaration of Helsinki. All subjects provided written informed consent. Reasons for exclusion of patients in this study were technical complications during surgery, lack of DNA, loss of follow-up, or re-transplantation at the time of recruitment. The primary endpoint of this study was long-term death-censored graft survival and the maximum follow-up period was 15 years. Graft failure was defined as the need for dialysis and/or re-transplantation. Secondary endpoints included: The occurrence of delayed graft function (DGF; described by the United Network for Organ Sharing as, “The need for at least one dialysis treatment in the first week after kidney transplantation,”) and biopsy-proven acute rejection (BPAR; based on the Banff ‘07 classification).

### DNA extraction and *MYH9* genotyping

Peripheral blood mononuclear cells from blood or splenocytes were obtained from both the donor and recipient. DNA isolation was done with a commercial kit according to the manufacturer’s instructions and stored at -80°C. Genotyping of the single nucleotide polymorphism (SNP) was performed using the Illumina VeraCode GoldenGate Assay kit as per the manufacturer’s instructions (Illumina, San Diego, CA, USA). We opted for the *MYH9* rs11089788 C>A SNP, which has previously been associated with kidney function in healthy individuals and with disease progression in patients with CKD.^13–15^ Genotype clustering and calling were performed using BeadStudio Software (Illumina). The overall genotype success rate was 99.9% and only two samples were excluded from subsequent analyses because of a missing call rate.

### Statistical analysis

SPSS software version 25 (SPSS Inc, Chicago, IL, USA) was used for our statistical analyses. Data are presented as the total number of patients with percentage [n (%)] for nominal variables, mean ± standard deviation for parametric variables, and median [IQR] for non-parametric variables. Differences among groups were tested with the Χ^2^ test for categorical variables or Student’s *t*-test for normally distributed variables, and the Mann-Whitney *U*-test for not-normally divided variables, respectively. The Log-rank test was used to test for differences in kidney allograft survival or rejection-free survival among the different genotypes. Logistic regression was used to assess the association of the *MYH9* polymorphism with DGF. Univariable analysis was used to examine the association of the *MYH9* polymorphism, recipient, donor, and transplant characteristics with BPAR as well as death-censored graft survival. Significant associations in univariable analyses were then assessed in a multivariable Cox regression. Two-tailed tests were regarded as significant at *P* < 0.05.

## Results

### Study population and determinants of graft failure

All patients who underwent a single kidney transplantation at the University Medical Center Groningen were recruited for this study (n=1,271). Baseline patient characteristics are shown in Table 1. During the mean study period of 6.2 years ± 4.2, 215 of 1,271 kidney transplant recipients (16.9%) developed graft failure. The main reason for graft failure was rejection (n = 126, containing acute rejection, transplant glomerulopathy, and chronic antibody-mediated rejection). Other causes for graft loss were surgical complications (n = 33), relapse of the original kidney disease (n = 16), other causes (n = 16), vascular disease (n = 12), and unknown cause (n = 12). In univariable analysis, DGF, recipient age, recipient blood type (AB *vs* others), donor type (living *vs* cadaveric), donor age, donor blood type (AB *vs* others), cold ischemia time, warm ischemia time, use of cyclosporin, and use of corticosteroids were all associated with graft failure (*P* < 0.05).

**Table 1:**
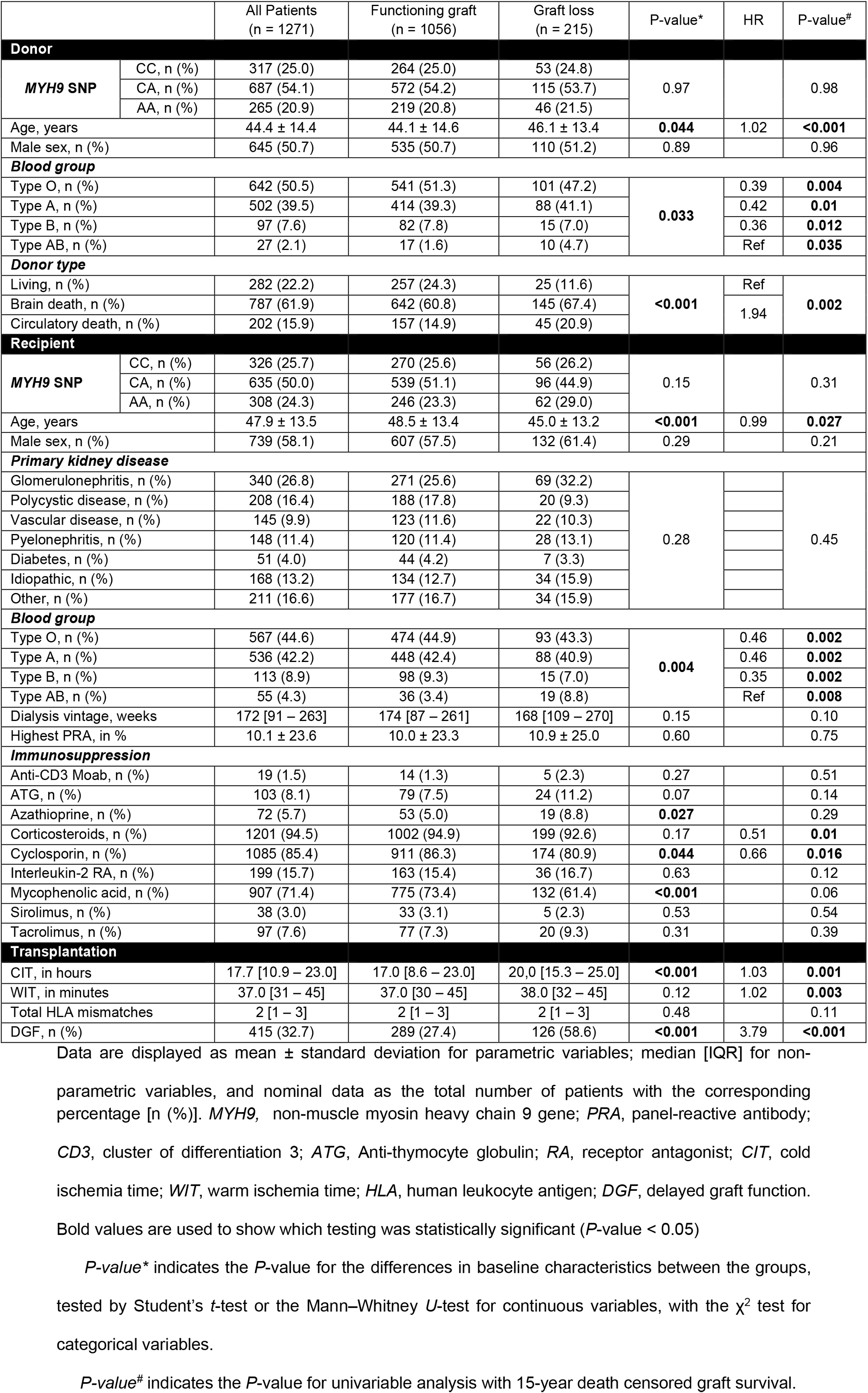
Baseline characteristics of the donors and recipients

|  |  | All Patients<br>(n = 1271) | Functioning graft<br>(n = 1056) | Graft loss<br>(n = 215) | P-value* | HR | P-value# |
| --- | --- | --- | --- | --- | --- | --- | --- |
| Donor |  |  |  |  |  |  |  |
| MYH9 SNP | CC, n (%) | 317 (25.0) | 264 (25.0) | 53 (24.8) | 0.97 |  | 0.98 |
|  | CA, n (%) | 687 (54.1) | 572 (54.2) | 115 (53.7) |  |  |  |
|  | AA, n (%) | 265 (20.9) | 219 (20.8) | 46 (21.5) |  |  |  |
| Age, years |  | 44.4 ± 14.4 | 44.1 ± 14.6 | 46.1 ± 13.4 | 0.044 | 1.02 | <0.001 |
| Male sex, n (%) |  | 645 (50.7) | 535 (50.7) | 110 (51.2) | 0.89 |  | 0.96 |
| Blood group |  |  |  |  |  |  |  |
| Type O, n (%) |  | 642 (50.5) | 541 (51.3) | 101 (47.2) | 0.033 | 0.39 | 0.004 |
| Type A, n (%) |  | 502 (39.5) | 414 (39.3) | 88 (41.1) |  | 0.42 | 0.01 |
| Type B, n (%) |  | 97 (7.6) | 82 (7.8) | 15 (7.0) |  | 0.36 | 0.012 |
| Type AB, n (%) |  | 27 (2.1) | 17 (1.6) | 10 (4.7) |  | Ref | 0.035 |
| Donor type |  |  |  |  |  |  |  |
| Living, n (%) |  | 282 (22.2) | 257 (24.3) | 25 (11.6) | <0.001 | Ref | 0.002 |
| Brain death, n (%) |  | 787 (61.9) | 642 (60.8) | 145 (67.4) |  | 1.94 |  |
| Circulatory death, n (%) |  | 202 (15.9) | 157 (14.9) | 45 (20.9) |  |  |  |
| Recipient |  |  |  |  |  |  |  |
| MYH9 SNP | CC, n (%) | 326 (25.7) | 270 (25.6) | 56 (26.2) | 0.15 |  | 0.31 |
|  | CA, n (%) | 635 (50.0) | 539 (51.1) | 96 (44.9) |  |  |  |
|  | AA, n (%) | 308 (24.3) | 246 (23.3) | 62 (29.0) |  |  |  |
| Age, years |  | 47.9 ± 13.5 | 48.5 ± 13.4 | 45.0 ± 13.2 | <0.001 | 0.99 | 0.027 |
| Male sex, n (%) |  | 739 (58.1) | 607 (57.5) | 132 (61.4) | 0.29 |  | 0.21 |
| Primary kidney disease |  |  |  |  |  |  |  |
| Glomerulonephritis, n (%) |  | 340 (26.8) | 271 (25.6) | 69 (32.2) | 0.28 |  | 0.45 |
| Polycystic disease, n (%) |  | 208 (16.4) | 188 (17.8) | 20 (9.3) |  |  |  |
| Vascular disease, n (%) |  | 145 (9.9) | 123 (11.6) | 22 (10.3) |  |  |  |
| Pyelonephritis, n (%) |  | 148 (11.4) | 120 (11.4) | 28 (13.1) |  |  |  |
| Diabetes, n (%) |  | 51 (4.0) | 44 (4.2) | 7 (3.3) |  |  |  |
| Idiopathic, n (%) |  | 168 (13.2) | 134 (12.7) | 34 (15.9) |  |  |  |
| Other, n (%) |  | 211 (16.6) | 177 (16.7) | 34 (15.9) |  |  |  |
| Blood group |  |  |  |  |  |  |  |
| Type O, n (%) |  | 567 (44.6) | 474 (44.9) | 93 (43.3) | 0.004 | 0.46 | 0.002 |
| Type A, n (%) |  | 536 (42.2) | 448 (42.4) | 88 (40.9) |  | 0.46 | 0.002 |
| Type B, n (%) |  | 113 (8.9) | 98 (9.3) | 15 (7.0) |  | 0.35 | 0.002 |
| Type AB, n (%) |  | 55 (4.3) | 36 (3.4) | 19 (8.8) |  | Ref | 0.008 |
| Dialysis vintage, weeks |  | 172 [91 – 263] | 174 [87 – 261] | 168 [109 – 270] | 0.15 |  | 0.10 |
| Highest PRA, in % |  | 10.1 ± 23.6 | 10.0 ± 23.3 | 10.9 ± 25.0 | 0.60 |  | 0.75 |
| Immunosuppression |  |  |  |  |  |  |  |
| Anti-CD3 Moab, n (%) |  | 19 (1.5) | 14 (1.3) | 5 (2.3) | 0.27 |  | 0.51 |
| ATG, n (%) |  | 103 (8.1) | 79 (7.5) | 24 (11.2) | 0.07 |  | 0.14 |
| Azathioprine, n (%) |  | 72 (5.7) | 53 (5.0) | 19 (8.8) | 0.027 |  | 0.29 |
| Corticosteroids, n (%) |  | 1201 (94.5) | 1002 (94.9) | 199 (92.6) | 0.17 | 0.51 | 0.01 |
| Cyclosporin, n (%) |  | 1085 (85.4) | 911 (86.3) | 174 (80.9) | 0.044 | 0.66 | 0.016 |
| Interleukin-2 RA, n (%) |  | 199 (15.7) | 163 (15.4) | 36 (16.7) | 0.63 |  | 0.12 |
| Mycophenolic acid, n (%) |  | 907 (71.4) | 775 (73.4) | 132 (61.4) | <0.001 |  | 0.06 |
| Sirolimus, n (%) |  | 38 (3.0) | 33 (3.1) | 5 (2.3) | 0.53 |  | 0.54 |
| Tacrolimus, n (%) |  | 97 (7.6) | 77 (7.3) | 20 (9.3) | 0.31 |  | 0.39 |
| Transplantation |  |  |  |  |  |  |  |
| CIT, in hours |  | 17.7 [10.9 – 23.0] | 17.0 [8.6 – 23.0] | 20.0 [15.3 – 25.0] | <0.001 | 1.03 | 0.001 |
| WIT, in minutes |  | 37.0 [31 – 45] | 37.0 [30 – 45] | 38.0 [32 – 45] | 0.12 | 1.02 | 0.003 |
| Total HLA mismatches |  | 2 [1 – 3] | 2 [1 – 3] | 2 [1 – 3] | 0.48 |  | 0.11 |
| DGF, n (%) |  | 415 (32.7) | 289 (27.4) | 126 (58.6) | <0.001 | 3.79 | <0.001 |

Data are displayed as mean ± standard deviation for parametric variables; median [IQR] for non-
parametric variables, and nominal data as the total number of patients with the corresponding
percentage [n (%)]. *MYH9*, non-muscle myosin heavy chain 9 gene; *PRA*, panel-reactive antibody; *CD3*, cluster of differentiation 3; *ATG*, Anti-thymocyte globulin; *RA*, receptor antagonist; *CIT*, cold ischemia time; *WIT*, warm ischemia time; *HLA*, human leukocyte antigen; *DGF*, delayed graft function.
Bold values are used to show which testing was statistically significant ( $P$ -value < 0.05)
*P*-value\* indicates the *P*-value for the differences in baseline characteristics between the groups, tested by Student's *t*-test or the Mann–Whitney *U*-test for continuous variables, with the $\chi^2$ test for categorical variables.
*P*-value# indicates the *P*-value for univariable analysis with 15-year death censored graft survival.

### Distribution of the *MYH9* polymorphism

The observed genotypic frequencies of the *MYH9* SNP (rs11089788 C>A) did not differ between donors (n = 1269; CC, 25.0%; CA, 54.1%; AA, 20.9%) and recipients (n = 1269; CC, 25.7%; CA, 50.0%; AA, 24.3%) (*P* = 0.07). The distribution of the SNP was in Hardy−Weinberg equilibrium. Compared to the 1000 genomes project, the genotypic frequencies of the *MYH9* polymorphism in recipients and donors were significantly different (*P* < 0.001).^22^ In both recipients and donors, the A-allele of the *MYH9* SNP was more prevalent than the reported allele and genotype frequencies in the 1000 genomes project. The percentage of kidney allografts with DGF significantly differed based on the recipient *MYH9* genotype (33.7% in CC, 29.6% in CA, 37.7% in AA, *P* = 0.041), but not for the donor *MYH9* genotype (*P* = 0.93). For further analysis, heterozygotes (CA) and homozygotes (CC)-genotypes were combined into one group (CA/CC). In logistic regression, recipients carrying the AA-genotype of the *MYH9* polymorphism had a significantly elevated risk of DGF (OR = 1.34 compared to CA/CC-genotypes; 95%-CI: 1.03 – 1.76; *P* = 0.031). In multivariable logistic regression, the AA-genotype of the *MYH9* polymorphism in recipients was no longer significantly associated with the occurrence of DGF (OR =1.26 compared to CA/CC-genotypes; 95%-CI: 0.92 – 1.72; *P* = 0.15, Table 2). There was no difference in the overall BPAR frequency among the *MYH9* genotypes in the donor (34.7% in CC, 33.0% in CA, 35.8% in AA, *P* = 0.69). In contrast, the distribution of the *MYH9* polymorphism in the recipient showed a trend towards a higher risk of BPAR (31.6% in CC, 32.4% in CA, 39.3% in AA, *P* = 0.068, Fig. 2A). A significant association was found with BPAR when the AA-genotype of the *MYH9* polymorphism in the recipient was compared to CA-and CC-genotypes (39.3% in AA versus 32.2% in CA/CC, *P* = 0.021, Fig. 2B). In multivariable Cox regression, the AA-genotype of the *MYH9* polymorphism in recipients was no longer significantly associated with the occurrence of BPAR (HR = 1.22 compared to CA/CC-genotypes; 95%-CI: 0.97 – 1.54; *P* = 0.10, Table 3). In summary, although the AA-genotype of the *MYH9* polymorphism in recipients was associated with DGF and BPAR, the significance was lost when correcting for potential confounders.

**Table 2:** Logistic regression analysis for the risk of delayed graft function after kidney transplantation

| Variables | P-value | Hazard Ratio |
| --- | --- | --- |
| *MYH9* rs111089788 SNP in the recipient<br>(AA versus CA/CC) | 0.15 | 1.26 (0.92 – 1.72) |
| Donor age<br>(in years) | <0.001 | 1.02 (1.01 – 1.03) |
| Donor gender<br>(male versus female) | 0.001 | 1.61 (1.22 – 2.13) |
| Donor type<br>(deceased versus living) | 0.001 | 31.61 (4.14 – 214.57) |
| Total HLA mismatches | 0.006 | 1.16 (1.04 – 1.30) |
| Dialysis vintage<br>(in years) | 0.007 | 1.08 (1.02 – 1.14) |
| Warm ischemia time<br>(in minutes) | 0.020 | 1.01 (1.00 – 1.03) |
| Recipient age<br>(in years) | 0.44 | 1.00 (0.99 – 1.02) |
| Cold ischemia time<br>(in minutes) | 0.47 | 1.00 (1.00 – 1.00) |

Multivariable logistic regression was performed for delayed graft function after kidney transplantation. Only variables with a $P < 0.05$ in the univariable analysis were included. Data are presented as an odds ratio with a 95% confidence interval (CI) and $P$ -value. Donor age, donor gender, donor type, total HLA mismatches, dialysis vintage, and warm ischemia time were significant, whereas the *MYH9* SNP (rs111089788) in the recipient, recipient age, and cold ischemia time were not.

**Figure 2.**
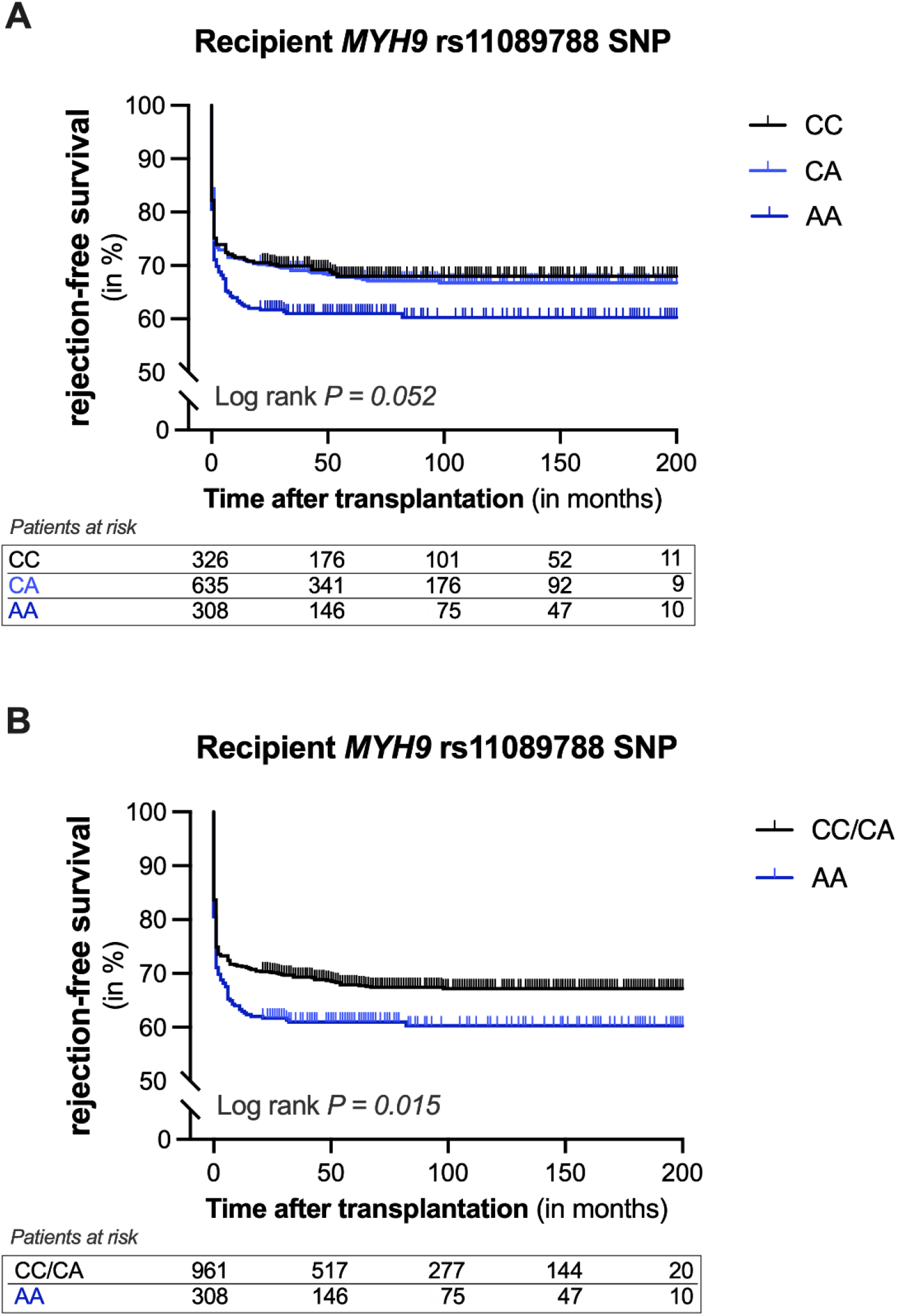
Kaplan-Meier curves for rejection-free survival of kidney allografts according to the presence of a non-muscle myosin heavy chain 9 gene polymorphism in the recipient. (**A**) Cumulative rejection-free survival of kidney allografts according to the presence of the *MYH9* SNP rs11089788 in the recipient. (**B**) Cumulative rejection-free survival of kidney allografts in recipients with the AA-genotype of the *MYH9* SNP rs11089788 versus the AC/CC-genotype. Log-rank test was used to compare the incidence of biopsy-proven rejection between the groups.

**Table 3:** Multivariable analysis for the risk of biopsy-proven acute rejection after kidney transplantation

| Variables | P-value | Hazard Ratio |
| --- | --- | --- |
| *MYH9* rs111089788 SNP in the recipient<br>(AA versus CA/CC) | 0.10 | 1.22 (0.97 – 1.54) |
| Recipient age<br>(in years) | <0.001 | 0.97 (0.97 – 0.98) |
| Total HLA mismatches | <0.001 | 1.20 (1.11 – 1.29) |
| Delayed graft function<br>(yes versus no) | 0.015 | 1.31 (1.05 – 1.62) |
| Recipient gender<br>(female versus male) | 0.039 | 1.25 (1.01 – 1.55) |
| Warm ischemia time<br>(in minutes) | 0.08 | 0.99 (0.98 – 1.00) |

Multivariable Cox regression was performed for biopsy-proven acute rejection (BPAR) after kidney transplantation. Only variables with a $P < 0.05$ in the univariable analysis were included. Data are presented as a hazard ratio with a 95% confidence interval (CI) and $P$ -value. Recipient age, total HLA mismatches, delayed graft function (DGF), and recipient gender were significant, whereas the *MYH9* SNP (rs111089788) in the recipient and warm ischemia time were not.

### Long-term kidney graft survival based on the *MYH9* genotypes

Kaplan–Meier survival analysis demonstrated no association between the *MYH9* SNP in the recipient or the donor and death-censored kidney graft survival (Figure 3). However, a trend was seen for a heightened rate of graft failure in the recipients with an AA-genotype of the *MYH9* polymorphism compared to CA- and CC-genotypes (graft loss: 33.2% in AA versus 24.1% in CA/CC, *P* = 0.056, Fig. 3B). Next, donor-recipient pairs were separated into four groups according to the presence or absence of the AA-genotype of the *MYH9* polymorphism in the donor and recipient. Kaplan–Meier survival analyses showed a significant difference in graft failure rates among the four groups (*P* = 0.036, Fig. 4A). Intriguingly, the AA-genotype of the *MYH9* polymorphism in the donor seemed to have a marginal positive impact on graft survival, whereas the AA-genotype in the recipient had a modest detrimental impact compared to donor-recipient pairs with the combined CC/CA-genotype. Recipients with an AA-genotype receiving a graft with an AA-genotype had the worst outcome. This combined genotype was identified in 6.3% of the donor-recipient pairs. Moreover, the significant association with graft failure increased when the combined AA-genotype of the *MYH9* polymorphism in donor-recipient pairs was compared to the other groups (*P* = 0.011, Fig. 4B). The cumulative 15-year death-censored kidney allograft survival was 50.4% in this combined AA-genotype group and 74.9% in the reference group, respectively. These data suggest that matching donor-recipient pairs on the *MYH9* polymorphism may impact long-term graft survival in kidney transplantation.

**Figure 3.**
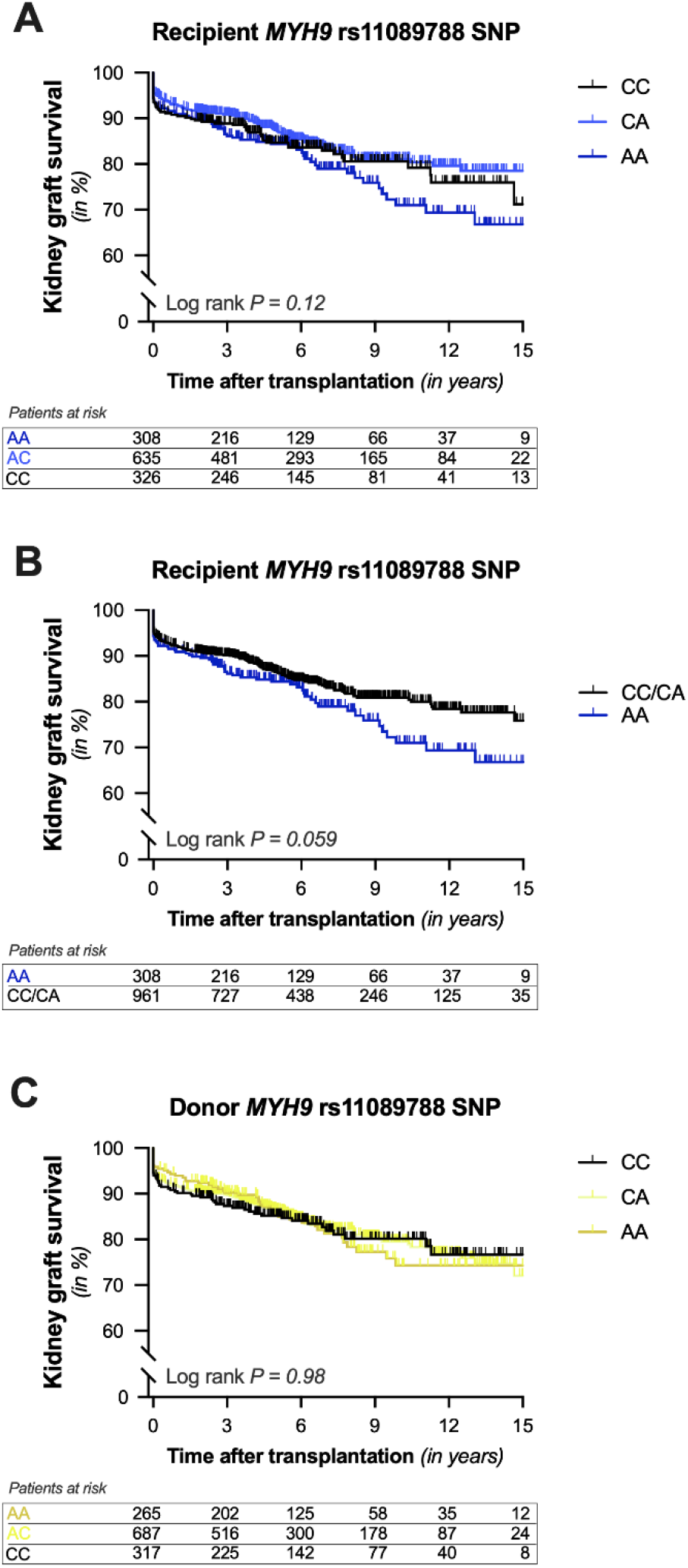
Kaplan-Meier curves for 15-year death-censored kidney graft survival according to the presence of a non-muscle myosin heavy chain 9 gene polymorphism in the donor or recipient. (**A**) Cumulative 15-year death-censored kidney graft survival according to the presence of a genetic variant in non-muscle myosin heavy chain 9 gene (*MYH9*, rs11089788 C>A) in (A, B) the recipient (blue line) or (C) the donor (yellow line). (**B**) Cumulative 15-year death-censored graft survival of kidney allografts in recipients with the AA-genotype of the *MYH9* SNP rs11089788 versus the AC/CC-genotype. Log-rank test was used to compare the incidence of graft loss between the groups.

**Figure 4.**
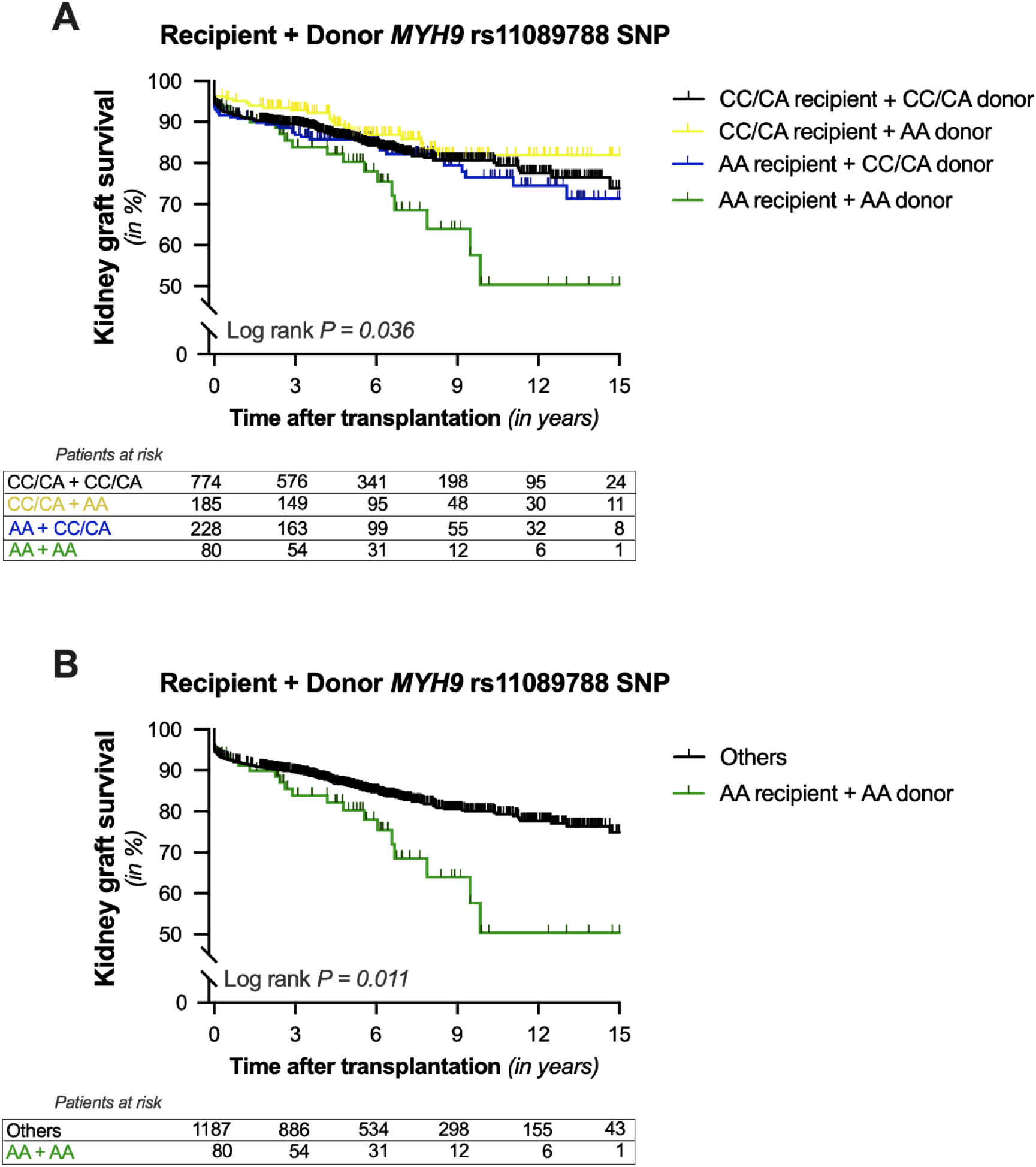
Kaplan-Meier curves for 15-year death-censored kidney graft survival according to the presence of a non-muscle myosin heavy chain 9 gene polymorphism in donor-recipient pairs. Cumulative 15-year death-censored kidney graft survival is shown according to the presence of the *MYH9* polymorphism in donor-recipient pairs. (**A**) Pairs were divided into four groups according to the absence (black line) or presence of the AA-genotype in the recipient (blue line), donor (yellow line), or both (green line). (**B**) In addition, the presence of the AA-genotype in both the recipient and donor (green line) was compared to the rest (black line). Log-rank test was used to compare the incidence of graft loss between the groups.

### Regression analysis for the *MYH9* polymorphism in donor-recipient pairs and graft failure

Finally, we investigated if the *MYH9* variant in donor-recipient pairs is an independent risk factor for graft failure. In univariable analysis, the combined AA-genotype of the *MYH9* SNP in donor-recipient pairs was associated with a hazard ratio of 1.78 (95%-CI: 1.13 – 2.79; *P* = 0.012) for graft failure after complete follow-up. Subsequently, multivariable analysis was performed to adjust for potential confounders, including donor and patient characteristics as well as transplant variables (Table 4). In these Cox regression analyses, the combined AA-genotype of the *MYH9* SNP in donor-recipient pairs remained significantly associated with graft failure. Lastly, we performed a multivariable Cox regression analysis using all variables that were significantly associated with graft failure in univariable analysis (Table 5). In this model, the *MYH9* SNP (rs11089788) in donor-recipient pairs, the occurrence of DGF, recipient age, and donor age were all significantly associated with graft loss. After adjustment, the hazard ratio for graft failure of the combined AA-genotype for the *MYH9* SNP in donor-recipient pairs was 1.68 (95% CI: 1.05 – 2.70, *P* = 0.031). In conclusion, our results reveal that recipients with an AA-genotype of the *MYH9* SNP receiving a kidney allograft with an AA-genotype have a significantly elevated risk of graft failure after kidney transplantation.

**Table 4:** Associations of non-muscle myosin heavy chain 9 polymorphism in donor-recipient pairs with graft loss after kidney transplantation.

|  | *MYH9* SNP (rs1800472) in donor-recipient pairs |  |  |
| --- | --- | --- | --- |
|  | Hazard ratio<br>(AA+AA vs. *others*) | 95% CI | *P*-value |
| **Model 1** | 1.78 | 1.13 – 2.79 | 0.012 |
| **Model 2** | 1.90 | 1.19 – 3.02 | 0.007 |
| **Model 3** | 1.95 | 1.24 – 3.08 | 0.004 |
| **Model 4** | 1.91 | 1.16 – 3.12 | 0.011 |

Data are presented as a hazard ratio with a 95% confidence interval (CI) and *P*-value.
**Model 1:** Crude model.
**Model 2:** Adjusted for model 1 plus recipient characteristics: Recipient age, recipient sex, recipient blood type, and dialysis vintage.
**Model 3:** Adjusted for model 1 plus donor characteristics: Donor age, donor sex, donor blood type, and donor origin.
**Model 4:** Adjusted for model 1 plus transplant characteristics: Cold and warm ischemia time, and the number of human leukocyte antigen (HLA)-mismatches.

**Table 5:** Multivariable analysis for the risk of graft loss after renal transplantation

| Variables | P-value | Hazard Ratio |
| --- | --- | --- |
| rs111089788 in donor-recipient pairs<br>(AA+AA versus *others*) | 0.031 | 1.68 (1.05 – 2.70) |
| Delayed graft function<br>(yes versus *no*) | <0.001 | 3.47 (2.56 – 4.72) |
| Recipient age<br>(in years) | <0.001 | 0.98 (0.97 – 0.99) |
| Donor age<br>(in years) | 0.001 | 1.02 (1.01 – 1.03) |
| Recipient blood type<br>(AB versus *other*) | 0.061 |  |
| Warm ischemia time<br>(in minutes) | 0.12 | 1.01 (1.00 – 1.02) |
| Corticosteroids | 0.20 | 1.53 (0.80 – 2.95) |
| Cold ischemia time<br>(in minutes) | 0.32 | 1.00 (1.00 – 1.00) |
| Donor type<br>(living versus *deceased*) | 0.41 | 0.76 (0.39 – 1.46) |
| Cyclosporin A | 0.71 | 1.04 (0.71 – 1.66) |
| Donor blood type<br>(AB versus *other*) | 0.90 |  |

Multivariable cox regression was performed for kidney graft survival. Only variables with a $P < 0.05$ in the univariable analysis were included. Data are presented as a hazard ratio with a 95% confidence interval (CI) and $P$ -value. In the final model, the *MYH9* SNP (rs11089788) in donor-recipient pairs, the occurrence of delayed graft function, recipient age, and donor age were significant, whereas recipient blood type, warm ischemia time, use of corticosteroids, cold ischemia time, donor type, use of cyclosporin A, and donor blood type were not.

## Discussion

A multitude of strategies can be pursued to improve the long-term outcomes after kidney transplantation, ranging from the development of novel drugs that can pull the brakes on allo-immune cascades, to the refinement of donor-recipient matching systems to minimize the severity of allograft recognition. Regarding allograft matching, HLA-centric systems remains the cornerstone of allocating kidney allografts—although a paradigm shift in the approach to donor-recipient matching is urgently needed.^23^ Genetic analyses in transplantation provides a particularly unique opportunity for the development of innovative strategies that can improve donor-recipient pairing and drive personalized medicine, in part by enabling individualized risk stratification.^24,25^ Presently, we report the impact of a recently discovered polymorphism in *MYH9* on long-term kidney allograft survival. The key finding of our study is that recipients with an AA-genotype of the *MYH9* rs11089788 variant receiving a kidney allograft with an AA-genotype of the same variant, have a significantly elevated risk of developing graft loss. In contrast, no association for the *MYH9* polymorphism with long-term allograft survival was found in either the recipient or donor when assessed individually. Hence, our study provides evidence that matching recipients with donor kidneys based on the *MYH9* polymorphism may well impact the risk of graft loss.

To our knowledge, our study is the first to show an association between this *MYH9* variant and long-term graft survival after kidney transplantation. Specifically, we found that the combined AA-genotype in donor-recipient pairs nearly doubled the risk of graft failure. Genome-wide linkage analysis recently highlighted the *MYH9* rs11089788 polymorphism as a top variant for kidney function in a meta-analysis of three European populations.^14^ In accordance with our results, the C-allele of the *MYH9* rs11089788 polymorphism was consistently associated with better kidney function in healthy Europeans.^14^ Furthermore, in a Chinese cohort of IgA nephropathy patients, the A-allele of this variant was associated with hastened progression to kidney failure.^13^ Other groups, however, did not recapitulate an association between this *MYH9* variant and kidney outcomes.^26,27^ In particular, Franceschini and colleagues found no relationship between the *MYH9* rs11089788 polymorphism and kidney function or CKD in native Americans.^27^ Importantly, we found no relationship of this *MYH9* variant in the recipient or the donor alone with death-censored kidney graft survival either. Our findings, thus, suggest that only donor-recipient interactions in *MYH9* may lead to kidney function decline after renal transplantation.

The importance of the *MYH9* for the kidney has been investigated by several group but remains controversial. Initial reports linked certain variants in the *MYH9* to a greater risk of CKD.^9,10^ Later studies uncovered that this association was based on the strong linkage disequilibrium between *MYH9* variants and variants in *APOL1*.^7,11^ Nonetheless, patients with rare mutations in *MYH9* leading to *MYH9*-related diseases often present with signs of CKD and can develop ESKD.^7,8^ In line with these results, heterozygous mice with mutations in *Myh9* manifest similar pathological kidney phenotypes as humans with *MYH9-*related diseases, including proteinuria, focal segmental glomerulosclerosis, and CKD.^28^ Intriguingly, *Myh9* knockdown in zebrafish lead to the malformation and dysfunction of their glomeruli.^29^ More specifically, these zebrafish failed to correctly develop the glomerular capillary structure, lacking fenestration in the endothelial cells and having an absence or reduced number of mesangial cells together with irregular thickening of the glomerular basement membrane.^29^ Although kidney clearance experiments showed that the glomerular barrier function remained unaltered, glomerular filtration in these zebrafish was significantly reduced.^29^ Altogether, these findings attest a key role for *MYH9* and non-muscle myosin heavy chain II-A in kidney development and physiology.

In humans, non-muscle myosin II-A, whose heavy chains are encoded by *MYH9*, is expressed in the podocytes, tubular cells, endothelial cells of the peritubular capillaries, interlobular arteries, and arterioles.^30^ A potential mechanism underpinning the association between *MYH9* polymorphism and graft failure would likely be dependent on kidney-expressed non-muscle myosin heavy chain II-A. On the basis of our findings, however, alternative mechanisms may be more probable. Firstly, in the recipients a trend was found for the association between the *MYH9* polymorphism and graft loss, while there was no association in the donor genotypes. Secondly, the AA-genotype of the *MYH9* variant in the recipient, but not the donor, was associated with BPAR and DGF, although significance was lost after adjusting for potential confounders. Lastly, in the genotypic analysis of the donor-recipient pairs, the isolated donor AA-genotype was marginally protective while the isolated AA-genotype in the recipient had a modest detrimental effect on graft survival. Additional evidence supporting a systemic role of the *MYH9* variant in determining kidney allograft outcomes is provided by case report of a patient with focal segmental glomerulosclerosis, where proteinuria rapidly recurred following a deceased donor kidney transplantation that therapeutically responded to plasmapheresis.^31^ Moreover, the fact that donor-recipient pairs with the combined AA-genotype of the *MYH9* variant had the highest risk of graft loss in our population, suggests both donor-recipient interactions in *MYH9* with perhaps a leading role for extra-renal expressed non-muscle myosin heavy chain II-A. A case report of two kidney transplants in pediatric patients suggested a similar donor-recipient *MYH9* interaction.^32^

Overall, there are several limitations to our study that warrant consideration. First, our study design is observational in nature and thus cannot determine whether associations are based on causality. Therefore, we cannot exclude the possibility that the *MYH9* rs11089788 variant is a tag SNP in the neighboring *APOL1* to *APOL6* region, justifying further investigation in this regard. Second, we investigated a single polymorphism in *MYH9* and did not examine the impact of *MYH9* haplotypes. Nevertheless, crucial strengths of our study were the analysis of the recently described *MYH9* polymorphism in both donors and recipients, our large patient population, the long and complete follow-up, and the hard clinical endpoints.

In conclusion, we found that patients with an AA-genotype of the *MYH9* rs11089788 variant receiving a donor kidney with the AA-genotype have an elevated risk of late graft loss. Considering the impact of this combined genotype, our findings imply donor-recipient interactions in *MYH9* that negatively influence the long-term allograft survival of kidney allografts.

## Data Availability

All data produced in the present study are available upon reasonable request to the authors.

## Abbreviations

*APOL1*: Apolipoprotein L1 gene
BPAR: Biopsy-proven acute rejection
CIT: Cold ischemia time
CKD: Chronic kidney disease
DBD: Donation after circulatory death
DCD: Donation after brain death
DGF: Delayed graft function
ESKD: End-stage kidney disease
HLA: Human leukocyte antigen
HR: Hazard ratio
*MYH9*: Myosin heavy chain 9 gene
PRA: Panel-reactive antibody
OR: Odds ratio
SNP: Single-nucleotide polymorphism
WIT: Warm ischemia time

## Acknowledgments

The authors thank the members of the REGaTTA cohort (REnal GeneTics TrAnsplantation; University Medical Center Groningen, University of Groningen, Groningen, the Netherlands): S. J. L. Bakker, J. van den Born, M. H. de Borst, H. van Goor, J. L. Hillebrands, B. G. Hepkema, G. J. Navis and H. Snieder. The illustrations of Figure 1 were made by Siawosh K. Eskandari.

